# Linking Adiponectin Gene Variants (+45T>G and +276G>T) to Adipokine Levels and Metabolic Syndrome in a North Indian Adult Women

**DOI:** 10.1101/2024.08.15.24311969

**Authors:** Abhishek Gupta, Arun Kumar Singh, Priyanka Gupta, Vani Gupta

**Affiliations:** Department of Physiology, King George’s Medical University, Lucknow-226003 (UP), India; Department of Medicine, King George’s Medical University, Lucknow-226003 (UP), India; Indian Institute of Public Health Gandhinagar, Gandhinagar-382042, Gujarat, India; Department of Biochemistry, College of Dental Science & Hospital, Amargadh, Bhavnagar, Gujarat, India

**Keywords:** adiponectin, gene variants, metabolic syndrome, adipokines, adult women

## Abstract

**Background:** Adiponectin, an adipocyte-derived adipokine, is often downregulated in obesity-related disorders. This study aimed to explore the association between adiponectin gene variants (+45T>G, rs2241766, and +276G>T, rs1501299) and circulating adipokine levels as well as metabolic syndrome in North Indian adult women.

**Methods:** We genotyped single nucleotide polymorphisms (SNPs) in 541 adult women, comprising 269 with metabolic syndrome (MetS) according to NCEP-ATP III criteria and 272 without MetS (wMetS; control). We assessed circulating levels of adiponectin, leptin, lipid profile, glucose, insulin, and HOMA-IR.

**Results:** Significant differences (p<0.01) were observed in circulating adipokines (adiponectin and leptin), lipid profile, glucose, insulin, HOMA-IR, and waist-to-hip ratio (WHR) between wMetS and MetS women. The frequency of the combined mutant genotype (TG+GG) at +45T>G was significantly lower (p=0.017) in MetS women, while the mutant G allele was higher (p=0.008) compared to the wild type. For the +276G>T variant, the frequency of the mutant T allele was significantly lower (p=0.027) in MetS women compared to wMetS women. The mutant genotypes GG of +45T>G and TT of +276G>T were significantly associated with lower adiponectin levels, higher leptin levels, and increased HOMA-IR (all p<0.001) in MetS women.

**Conclusions:** The findings suggest that adiponectin gene variants (+45T>G and +276G>T), along with reduced adiponectin levels and elevated HOMA-IR, may contribute significantly to the development of metabolic syndrome.

## Introduction

Metabolic syndrome is a prevalent metabolic disorder characterized by a cluster of risk factors, including increased waist circumference, elevated triglyceride levels, low HDL-cholesterol, hyperglycemia, and hypertension **(Gupta A et al., 2010)**. Central adiposity, a key component of metabolic syndrome, is more common in women than in men **(Lorenzo C et al., 2003)**, largely due to the strong correlation between waist circumference and increased adiposity.

Adipose tissue, a crucial endocrine organ, plays a significant role in regulating whole-body metabolism and other essential functions related to inflammation and immune responses. There is considerable interest in understanding how adipose tissue-derived adipokines might mediate the relationship between body fat distribution and insulin sensitivity. Adiponectin, one of the most studied adipokines, is an insulin-sensitizing hormone, also known as APM1 or adipoQ. This adipose tissue-specific protein consists of 247 amino acids and plays a crucial role in energy homeostasis and insulin sensitivity **(Nguyen TMD, 2020)**. It has anti-inflammatory properties that affect the NF-κB pathway and enhances insulin action on peripheral tissues. Another significant adipokine, leptin, acts as an “adiposity signal” that modulates appetite and maintains energy balance **(Bjorbaek C et al., 2004)**, potentially contributing to the development of metabolic syndrome. In humans, obesity is often associated with increased leptin levels and decreased adiponectin levels. Adiponectin is located on chromosome 3q27, a region identified as a susceptibility locus for metabolic syndrome, type 2 diabetes, and coronary artery disease in a genome-wide scan study **(Mohammadzadeh G et al., 2016)**. The most commonly studied SNP variants of adiponectin are located at positions +45T/G and +276G/T, present on exon 2 and intron 2, respectively.

Numerous studies have explored the association between these variants and circulating adiponectin levels, visceral obesity, insulin resistance syndrome, and metabolic risk factors **(Ukkola O et al.,2003; Palit SP et al., 2020; Jang Y et al.,2008)** in humans. However, the association between these adiponectin SNPs and metabolic syndrome has not been extensively studied in age-matched adult women from North India.

This study aims to investigate the association of metabolic risk and adipokine gene variants in adult women from this diverse region in India. Specifically, the study focuses on: (i) the frequency distribution of adiponectin gene variants (+45 T/G and +276 G/T); (ii) the impact of these variants on phenotypic, clinical, and biochemical profiles; (iii) the association of adiponectin variants with circulating adipokine levels, HOMA-IR, and metabolic risk factors; and (iv) the relationship between adiponectin SNPs, HOMA-IR, circulating adipokine levels, and the presence of metabolic syndrome and associated risk factors. Thus, we examined the genotypic variability of adiponectin and its association with metabolic syndrome in adult women.

## Materials and Methods

### Study population

This case-control study was conducted at King George’s Medical University (KGMU) in Lucknow, India. A total of 541 women aged 20-40 years were enrolled from the KGMU outpatient department and the general population of Uttar Pradesh, a northern region of India. The study participants were divided into two groups: 269 women with metabolic syndrome (MetS) (mean age 31.91 ± 6.05) based on **NCEP-ATP III criteria 2001**, and 272 age-matched healthy women without MetS (wMetS) (mean age 30.96 ± 7.01), who were non-alcoholic, non-diabetic, and free of cardiac, respiratory, inflammatory, endocrine, or metabolic diseases. Women who were pregnant, lactating, had gynecological or obstetrical issues, or were on medications including hormone replacement therapy were excluded from the study. A structured questionnaire collected information on medical, personal, family, dietary, and menstrual history. Institutional ethics committee of KGMU, Lucknow approved (Reference code:-XXI ECM/P7, no. 1854-R.Cell-06-07) the study and and “all applicable institutional and governmental regulations concerning the ethical use of human volunteers were followed during this research”. Written informed consent for the participation in the study was obtained prior to enrollment from all the participant women.

### Study Material and Laboratory Measurements

Each participant was assessed for body mass index (BMI), height, weight, waist circumference (WC), hip circumference (HC), and waist-to-hip ratio (WHR). BMI was calculated as body weight (kg) divided by height (m^2^), and WHR was used to measure central obesity by dividing WC (measured at the narrowest point above the hip) by HC (measured at the greatest gluteal protrusion). Women were classified as having MetS if they met three or more of the NCEP-ATP III criteria: central obesity (WC >88 cm), hypertension (systolic blood pressure >130 mm Hg or diastolic blood pressure >85 mm Hg), hypertriglyceridemia (triglycerides >150 mg/dl), low HDL-cholesterol (<50 mg/dl), or fasting plasma glucose >110 mg/dl.

Venous blood samples were collected in the morning after an overnight fast on the 10th day of menstruation. From 5 ml of blood, plasma and serum samples were either analyzed immediately or stored at −80°C. Commercial enzymatic test kits were used to determine blood plasma glucose and serum lipid profile by GOD-POD and enzymatic methods, respectively (Randox Laboratories Ltd., Antrim, UK). Adiponectin (R&D Systems Inc., Minneapolis, USA; sensitivity 0.25 ng/ml, intra-assay coefficient of variation 3.4%) and leptin (Diagnostics Biochem Canada Inc., Canada; sensitivity 0.50 ng/ml, intra-assay coefficient of variation 4.3%) levels were measured using sandwich enzyme-linked immunosorbent assay (ELISA) methods, and fasting plasma insulin was assessed by immunoradiometric assay (Immunotech Radiova, Prague). Insulin resistance was calculated using the homeostatic model assessment (HOMA) **(Matthews DR et al.,1985)** formula: [FPG (mmol/l) x fasting insulin (μU/ml)] / 22.5.

### Genotyping of Adiponectin Gene (+45 T/G and +276 G/T)

Genomic DNA was extracted from 3 ml of venous blood collected in an EDTA vial using a commercial genomic DNA purification kit (Qiagen, Valencia, CA, USA). Genotyping of adiponectin was performed using polymerase chain reaction (PCR) on a Thermo Cycler Instrument (Bio-Rad Inc., Hercules, CA, USA), followed by restriction fragment length polymorphism (RFLP) analysis. For adiponectin +45T/G, the forward primer was 5’-TCCTTTGTAGGTCCCAACT-3’ and the reverse primer was 5’-GCAGCAAAGCCAAAGTCTTG-3’. The PCR conditions were: 95°C for 5 min, 35 cycles of 45 s at 94°C, 45 s at 57.5°C, and 45 s at 72°C, followed by a 7 min extension at 72°C. The 503 bp PCR fragment was digested with BspH1/Pag1 at 37°C. For adiponectin +276G/T, the forward primer was 5’-AGAAAGCAGCTCCTAGAAGT-3’ and the reverse primer was 5’-GGCACCATCTACACTCATCC-3’. The PCR conditions were: 95°C for 4 min, 35 cycles of 45 s at 94°C, 45 s at 57.5°C, and 45 s at 72°C, followed by a 7 min extension at 72°C. The 518 bp fragment was typed using BgL1 at 37°C. Each PCR reaction was performed in a total volume of 25 μl containing 3-3.5 mm/l MgCl2, 0.5 mm of each dNTP (Bangalore Genei, India), 0.2 μm of each primer, 2 U of Taq DNA polymerase (Bangalore Genei, India), and 10 ng of genomic DNA. The genotyping products were separated by electrophoresis on 2% (w/v) agarose gels and visualized by ethidium bromide staining.

### Statistical Analysis

Genotype and allele distributions were compared between wMetS and MetS women using the Chi-square test (χ^2^). Hardy-Weinberg equilibrium was assessed using the Chi-square test. Continuous data were compared using the two-sample Student’s t-test, while categorical data were analyzed using the Chi-square test with Yates’s correction. Pearson correlation (r) was used to examine associations between genetic polymorphisms and demographic and biochemical variables. Multiple logistic regression analysis was used to estimate the association of adiponectin genes with MetS risk and its components, with a p-value <0.05 considered statistically significant. GraphPad Prism (version 9.0) software was used for analysis.

## Results

In this study, a total of 541 adult women (272 wMetS and 269 with MetS) were screened to investigate the frequency of adiponectin gene variants at positions +45T/G and +276G/T. An independent samples t-test was conducted to compare demographic, clinical, and biochemical markers between women with and without MetS (Table 1, Figure 1). Although, the women in both groups were age-matched, there was no significant difference in mean age (p > 0.05). Significant differences (p < 0.001) were found between the two groups in adiponectin and leptin levels, anthropometric variables, lipid profiles, glucose, insulin, and HOMA-IR.

**Table 1.** Comparison of demographic and biochemical variables between women with metabolic syndrome (MetS) and those without (wMetS).

| Markers | wMetS<br>(n=272) | MetS<br>(n=269) | P value |
| --- | --- | --- | --- |
| Age (yrs) | 30.96 ± 7.01 | 31.91 ± 6.05 | NS |
| **<u>Anthropometric variables</u>** |  |  |  |
| WC (cm) | 74.11 ± 10.44 | 88.71 ± 13.99 | <0.0001 |
| WHR | 0.82 ± 0.05 | 0.87 ± 0.05 | <0.0001 |
| BMI (Kg/m<sup>2</sup>) | 22.98 ± 4.36 | 28.10 ± 5.09 | <0.0001 |
| **<u>Blood Pressure</u>** |  |  |  |
| SBP (mm Hg) | 118.36 ± 8.82 | 132.20 ± 11.97 | <0.0001 |
| DBP (mm Hg) | 80.25 ± 6.50 | 88.17 ± 7.80 | <0.0001 |
| **<u>Lipid Profile</u>** |  |  |  |
| TC (mg/dl) | 147.33 ± 28.49 | 171.80 ± 33.9 | <0.0001 |
| HDL (mg/dl) | 43.75 ± 6.46 | 41.73 ± 4.85 | <0.0001 |
| TG (mg/dl) | 106.86 ± 23.15 | 143.20 ± 38.31 | <0.0001 |
| LDL (mg/dl) | 101.60 ± 31.46 | 82.20 ± 27.26 | <0.0001 |
| VLDL (mg/dl) | 21.36 ± 4.62 | 28.64 ± 7.66 | <0.0001 |
| **<u>Glucose homeostasis</u>** |  |  |  |
| FPG (mg/dl) | 91.08 ± 10.40 | 105.01 ± 17.68 | <0.0001 |
| FPI (μU/ml) | 8.48 ± 5.79 | 12.24 ± 8.42 | <0.0001 |
| HOMA-IR | 1.93 ± 1.39 | 3.31 ± 2.58 | <0.0001 |
| **<u>Adipokine levels</u>** |  |  |  |
| Adiponectin (ng/ml) | 29.56 ± 13.46 | 20.21 ± 10.85 | <0.0001 |
| Leptin (ng/ml) | 9.56 ± 6.88 | 15.77 ± 10.47 | <0.0001 |

Values are expressed as mean ± SD and significance (p <0.05) was calculated between wMetS vs MetS.
MetS, metabolic syndrome; wMetS, without metabolic syndrome; WC, waist circumference; WHR, waist to hip ratio; BMI, body mass index; SBP, systolic blood pressure; DBP, diastolic blood pressure; TC, total cholesterol; HDL, high density lipoprotein; TG, triglyceride; LDL, low density lipoprotein; VLDL, very low-density lipoprotein; FPG, fasting plasma glucose; FPI, fasting plasma insulin; HOMA-IR, homeostatic model assessment-insulin resistance; NS, Nonsignificant.

**Figure 1.**
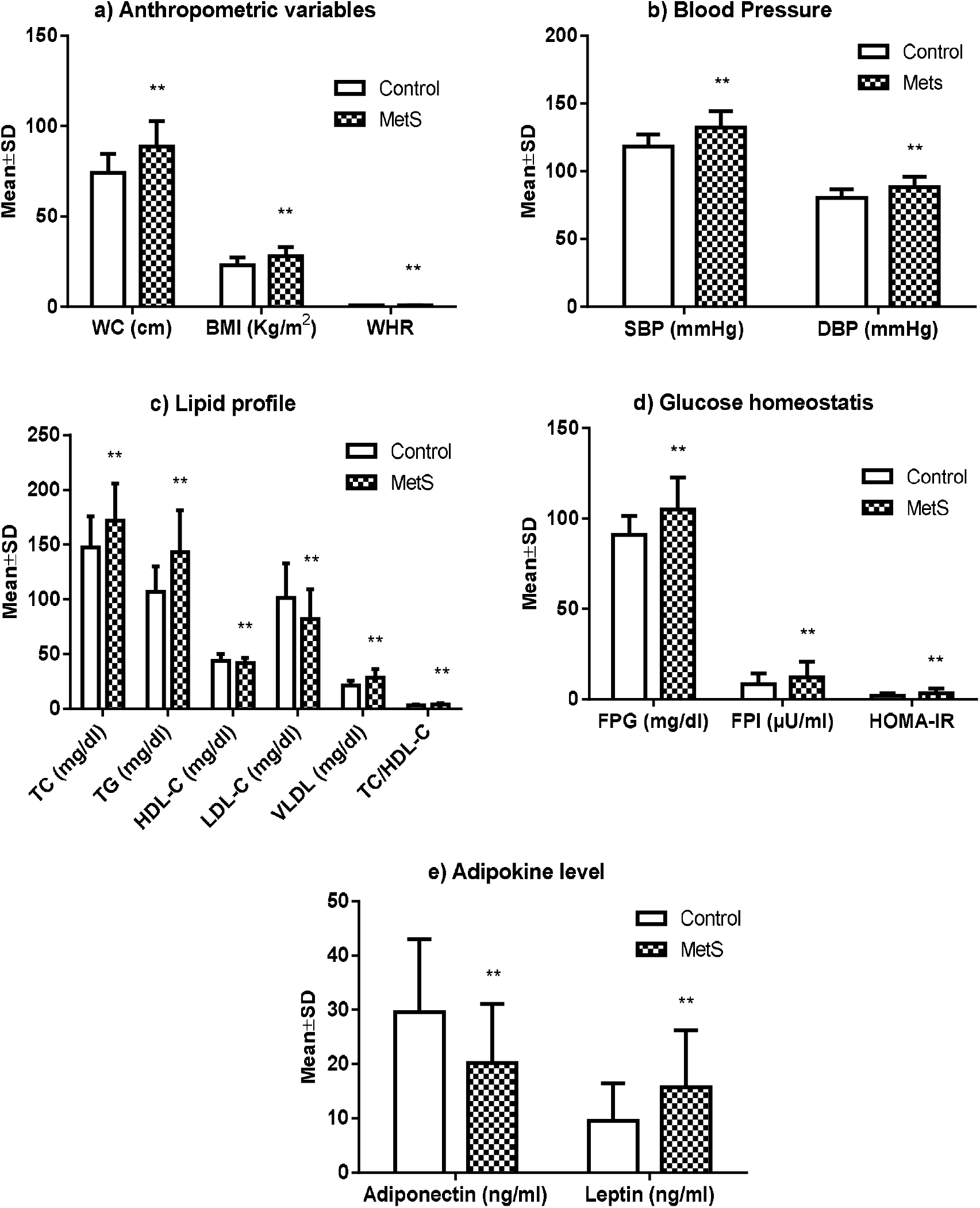
Anthropometric variables, risk factors, and circulating adiponectin and leptin levels in North Indian women with and without metabolic syndrome. MetS, metabolic syndrome; WC, waist circumference; BMI, body mass index; WHR, waist to hip ratio; SBP, systolic blood pressure; DBP, diastolic blood pressure; TC, total cholesterol; TG, triglyceride; HDL-C, high density lipoprotein-cholesterol; LDL-C, low density lipoprotein-cholesterol; VLDL, very low density lipoprotein; FPG, fasting plasma glucose; FPI, fasting plasma insulin; HOMA-IR, homeostasis model assessment-insulin resistance

Table 2 presents the frequencies of the adiponectin gene variants at positions +45 T/G and+276 G/T in the MetS and wMetS groups. The genotype distributions for both SNPs, +45 T/G (χ^2^ = 0.088; p = 0.767) and +276 G/T (χ^2^ = 0.157; p = 0.692), were in Hardy–Weinberg equilibrium. The frequency of the combined mutant TG+GG genotype at +45T/G (χ^2^ = 5.707, p < 0.05) and the mutant G allele (χ^2^ = 7.030, p = 0.008) differed significantly between the MetS and wMetS groups, with notable significance for the TG and GG genotypes. For adiponectin +276 G/T, only the mutant T allele (χ^2^ = 4.840, p = 0.028) showed a significant difference between the two groups, while genotype frequencies did not differ significantly (p > 0.05).

**Table 2.** Frequencies of adiponectin genotypes (+45T/G and +276G/T) in women with and without metabolic syndrome.

| SNPs | Genotype/<br>Alleles | wMetS<br>n=272 (%) | MetS<br>n=269 (%) | OR<br>(95% CI) | p value |
| --- | --- | --- | --- | --- | --- |
| +45 T/G | TT<sup>δ</sup> | 178 (65.4) | 148 (55.0) | Ref |  |
|  | TG | 85 (31.3) | 103 (38.3) | 1.46 (1.02-2.09) | 0.050 |
|  | GG | 9 (3.3) | 18 (6.7) | 2.41 (1.05-5.51) | 0.054 |
|  | TG+GG | 94 (34.6) | 121 (45.0) | 1.55 (1.10-2.91) | 0.017* |
|  | T<sup>δδ</sup> | 441 (81.1) | 399 (74.2) | Ref |  |
|  | G | 103 (18.9) | 139 (25.8) | 1.49 (1.12-1.99) | 0.008** |
| +276<br>G/T | GG<sup>δ</sup> | 152 (55.9) | 128 (47.6) | Ref |  |
|  | GT | 101 (37.1) | 111 (41.3) | 1.31 (0.91-1.87) | 0.171 |
|  | TT | 19 (7.0) | 30 (11.2) | 1.88 (1.01-3.49) | 0.064 |
|  | GT+TT | 120 (44.1) | 141 (52.4) | 1.40 (0.99-1.96) | 0.065 |
|  | G<sup>δδ</sup> | 405 (74.5) | 367 (68.2) | Ref |  |
|  | T | 139 (25.6) | 171 (31.8) | 1.36 (1.04-1.77) | 0.028* |

P values showed as <0.05\* and <0.01\*\* (Significant).
Wild genotype<sup>δ</sup> and wild allele<sup>δδ</sup> (taken as reference) compared with respective genotype and allele MetS, metabolic syndrome; wMetS, without metabolic syndrome; SNPs, Single nucleotide polymorphisms; OR, Odd ratio; CI, Confidence Interval

Table 3 analyzes the association between adiponectin variants and demographic and bio-clinical variables in the MetS group. The results indicate that waist-to-hip ratio (WHR) and adiponectin levels were significantly associated with +45T/G frequency (p < 0.05 and p < 0.01, respectively). However, no associations were found with other parameters in the wMetS group (data not shown). On the other hand, the adiponectin +276G/T variant was strongly associated with phenotypic variables such as waist circumference (WC), WHR, BMI, blood pressure (BP), and circulating leptin levels, except for adiponectin levels, lipid profiles, glucose, insulin, and HOMA-IR. In the wMetS group, only WC and HDL-C were significantly associated with the +276G/T variant (data not shown).

**Table 3.** Distribution of adiponectin gene variants (+45T/G and +276G/T) among women with metabolic syndrome (n=269).

| **+45 T/G** | **TT**<br>(n=148) | **TG**<br>(n=103) | **GG**<br>(n=18) | **P value** |
| --- | --- | --- | --- | --- |
| WC (cm) | 87.53 ± 13.47 | 89.54 ± 14.50 | 93.72 ± 14.51 | NS |
| WHR | 0.87 ± 0.05 | 0.88 ± 0.05 | 0.91 ± 0.04 | 0.004 |
| BMI (kg/m<sup>2</sup>) | 27.51 ± 5.04 | 28.73 ± 5.23 | 29.39 ± 4.33 | NS |
| SBP (mmHg) | 131.41 ± 12.47 | 132.80 ± 11.92 | 135.67 ± 6.41 | NS |
| DBP (mmHg) | 87.35 ± 7.55 | 88.76 ± 7.98 | 91.56 ± 8.67 | NS |
| TC (mg/dl) | 171.10 ± 34.99 | 173.18 ± 33.48 | 170.29 ± 29.18 | NS |
| TG (mg/dl) | 143.02 ± 37.84 | 145.13 ± 40.48 | 133.68 ± 28.42 | NS |
| HDL-C (mg/dl) | 41.82 ± 4.91 | 41.44 ± 4.72 | 42.61 ± 5.26 | NS |
| TC/HDL-C | 4.14 ± 0.96 | 4.24 ± 0.98 | 4.06 ± 0.92 | NS |
| FPG (mg/dl) | 104.23 ± 17.25 | 106.48 ± 18.05 | 103.07 ± 19.55 | NS |
| Insulin (μU/ml) | 12.06 ± 8.01 | 12.41 ± 8.96 | 12.89 ± 9.09 | NS |
| HOMA-IR | 3.25 ± 2.48 | 3.37 ± 2.63 | 3.61 ± 3.22 | NS |
| Adiponectin (ng/ml) | 21.39 ± 10.65 | 19.73 ± 11.76 | 13.20 ± 5.91 | 0.0096 |
| Leptin (ng/ml) | 15.62 ± 10.56 | 15.73 ± 9.32 | 17.28 ± 15.56 | NS |

| **+276 G/T** | **GG**<br>(n=128) | **GT**<br>(n=111) | **TT**<br>(n=30) | **P value** |
| --- | --- | --- | --- | --- |
| WC (cm) | 87.30 ± 13.38 | 88.82 ± 14.51 | 94.33 ± 13.58 | 0.046 |
| WHR | 0.87 ± 0.05 | 0.87 ± 0.05 | 0.91 ± 0.05 | 0.0003 |
| BMI (kg/m<sup>2</sup>) | 27.38 ± 4.65 | 28.23 ± 5.28 | 30.70 ± 5.49 | 0.005 |
| SBP (mmHg) | 130.27 ± 13.54 | 134.34 ± 10.52 | 132.77 ± 8.23 | 0.030 |
| DBP (mmHg) | 86.69 ± 7.95 | 89.78 ± 7.33 | 88.53 ± 8.30 | 0.009 |
| TC (mg/dl) | 169.14 ± 33.65 | 173.11 ± 36.70 | 178.65 ± 22.47 | NS |
| TG (mg/dl) | 140.96 ± 34.85 | 145.99 ± 44.42 | 142.51 ± 26.63 | NS |
| HDL-C (mg/dl) | 42.05 ± 4.71 | 41.31 ± 5.03 | 41.92 ± 4.82 | NS |
| TC/HDL-C | 4.07 ± 0.92 | 4.25 ± 1.06 | 4.32 ± 0.75 | NS |
| FPG (mg/dl) | 107.04 ± 18.34 | 103.39 ± 17.50 | 102.37 ± 14.85 | NS |
| Insulin (μU/ml) | 11.88 ± 7.80 | 12.54 ± 9.26 | 12.74 ± 7.97 | NS |
| HOMA-IR | 3.28 ± 2.49 | 3.36 ± 2.77 | 3.35 ± 2.34 | NS |
| Adiponectin (ng/ml) | 20.99 ± 10.64 | 20.22 ± 10.82 | 16.84 ± 11.57 | NS |
| Leptin (ng/ml) | 14.75 ± 9.35 | 15.60 ± 9.64 | 20.75 ± 15.77 | 0.018 |

All values are expressed in mean±SD
Significant p values <0.05 (bold) and p >0.05= Nonsignificant (NS).
One way ANOVA analysis, n=269 (metabolic syndrome women)

In Table 4 (a & b), a multiple regression analysis was performed to identify the strongest associations between adiponectin +45G and +276T mutant alleles and various variables across all women combined. The independent variables included BMI, WC, WHR, HDL-C, TG, LDL-C, TC/HDL-C, fasting glucose, insulin, insulin resistance, and leptin levels. The multivariate regression analysis revealed that WHR and adiponectin were the strongest and most significant predictors associated with the +45 mutant G allele, while glucose and systolic blood pressure (SBP) were strongly associated with the +276 mutant T allele.

**Table 4.** Associations between predictor variables in a total 541 adult women of North India:

a) Presence or absence of the mutant allele (G) of adiponectin +45 T/G gene
| Predictors | Coef | SE | t ratio | 95% CI |  | P value |
| --- | --- | --- | --- | --- | --- | --- |
|  |  |  |  | Lower | Upper |  |
| Constant | 0.502 | 0.205 | 2.451 | 0.101 | 0.904 | 0.015 |
| WC | -0.014 | 0.062 | 0.232 | -0.136 | 0.108 | NS |
| WHR | 0.242 | 0.117 | 2.065 | 0.012 | 0.471 | 0.039 |
| BMI | -0.076 | 0.121 | 0.627 | -0.313 | 0.161 | NS |
| Glucose | 0.007 | 0.059 | 0.113 | -0.109 | 0.123 | NS |
| SBP | 0.105 | 0.068 | 1.528 | -0.029 | 0.239 | NS |
| DBP | 0.003 | 0.065 | 0.051 | -0.124 | 0.130 | NS |
| TC | 0.004 | 0.119 | 0.037 | -0.228 | 0.237 | NS |
| TG | -0.107 | 0.291 | 0.369 | -0.678 | 0.463 | NS |
| HDL-C | 0.041 | 0.084 | 0.490 | -0.124 | 0.207 | NS |
| LDL-C | -0.013 | 0.091 | 0.143 | -0.191 | 0.165 | NS |
| TC/HDL-C | 0.022 | 0.052 | 0.428 | -0.079 | 0.123 | NS |
| Insulin | -0.041 | 0.099 | 0.412 | -0.234 | 0.153 | NS |
| HOMA-IR | 0.045 | 0.062 | 0.723 | -0.076 | 0.165 | NS |
| Adiponectin | 0.301 | 0.112 | 2.711 | 0.083 | 0.518 | 0.007 |
| Leptin | -0.076 | 0.054 | 1.392 | -0.183 | 0.031 | NS |

| Predictors | Coef | SE | t ratio | 95% CI |  | p value |
| --- | --- | --- | --- | --- | --- | --- |
|  |  |  |  | Lower | Upper |  |
| Constant | 0.455 | 0.212 | 2.141 | 0.038 | 0.871 | 0.033 |
| WC | 0.036 | 0.064 | 0.566 | -0.090 | 0.163 | NS |
| WHR | 0.007 | 0.121 | 0.055 | -0.231 | 0.245 | NS |
| BMI | 0.037 | 0.125 | 0.293 | -0.209 | 0.282 | NS |
| Glucose | -0.133 | 0.061 | 2.170 | -0.254 | -0.013 | 0.030 |
| SBP | 0.178 | 0.071 | 2.505 | 0.039 | 0.317 | 0.013 |
| DBP | -0.100 | 0.067 | 1.491 | -0.232 | 0.032 | NS |
| TC | -0.054 | 0.123 | 0.440 | -0.295 | 0.187 | NS |
| TG | -0.434 | 0.302 | 1.437 | -1.026 | 0.158 | NS |
| HDL-C | -0.041 | 0.088 | 0.470 | -0.213 | 0.130 | NS |
| LDL-C | 0.048 | 0.094 | 0.510 | -0.136 | 0.232 | NS |
| TC/HDL-C | 0.056 | 0.054 | 1.037 | -0.049 | 0.161 | NS |
| Insulin | -0.029 | 0.102 | 0.286 | -0.230 | 0.171 | NS |
| HOMA-IR | 0.115 | 0.064 | 1.803 | -0.010 | 0.240 | NS |
| Adiponectin | -0.038 | 0.115 | 0.334 | -0.264 | 0.187 | NS |
| Leptin | -0.078 | 0.056 | 1.380 | -0.189 | 0.033 | NS |

Values transformed in 0 (GG, n=280) and 1 (GT +TT, n=261)
Significant p values <0.05 (bold) and p >0.05= Nonsignificant (NS).
WC, waist circumference; WHR, waist to hip ratio; BMI, body mass index; SBP, systolic blood pressure; DBP, diastolic blood pressure; TG, triglyceride; HDL-C, high density lipoprotein-cholesterol; TC, total cholesterol; LDL-C, low density lipoprotein-Cholesterol; HOMA-IR, homeostasis model assessment-insulin resistance; Coef, coefficient; SE, standard error; 95 %CI, 95 % confidence interval

Table 5 illustrates the relationship between adiponectin and metabolic risk factors (Figure 2 & 3). Overall, adiponectin was inversely related to adiposity, fasting glucose, HOMA-IR, and leptin, and positively associated with HDL-C. In women with MetS, adiponectin showed an inverse correlation with WC, WHR, BMI, and leptin, and a positive correlation with HDL-C, while no significant associations were observed with other metabolic risk factors.

**Table 5.** Pearson correlation (r) of circulating adiponectin levels with anthropometric and biochemical variables in North Indian adult women.

| Variable | Overall<br>(N=541) |  | wMetS<br>(N=272) |  | MetS<br>(N=269) |  |
| --- | --- | --- | --- | --- | --- | --- |
|  | r | p-value | r | p-value | r | p-value |
| WC, cm | -0.5201 | <0.0001 | -0.4429 | <0.0001 | -0.4256 | <0.0001 |
| WHR | -0.4804 | <0.0001 | -0.3842 | <0.0001 | -0.4075 | <0.0001 |
| BMI, Kg/m<sup>2</sup> | -0.593 | <0.0001 | -0.5168 | <0.0001 | -0.531 | <0.0001 |
| SBP mm Hg | -0.226 | <0.0001 | -0.0459 | NS | -0.03099 | NS |
| DBP, mm Hg | -0.1957 | <0.0001 | -0.03619 | NS | -0.0225 | NS |
| TC, mg/dl | -0.2267 | <0.0001 | -0.1716 | 0.0045 | -0.05068 | NS |
| TG, mg/dl | -0.243 | <0.0001 | -0.1346 | 0.0264 | -0.04657 | NS |
| HDL-C, mg/dl | 0.2012 | <0.0001 | 0.1959 | 0.0012 | 0.07495 | NS |
| LDL-C, mg/dl | -0.2306 | <0.0001 | -0.2029 | 0.0008 | -0.0624 | NS |
| TC/HDL-C ratio | -0.2888 | <0.0001 | -0.27 | <0.0001 | -0.08268 | NS |
| FPG, mg/dl | -0.1887 | <0.0001 | -0.05096 | NS | -0.03642 | NS |
| FPI, μU/ml | -0.1334 | 0.0019 | -0.04603 | NS | -0.05276 | NS |
| HOMA-IR | -0.1637 | 0.0001 | -0.04283 | NS | -0.07436 | NS |
| Leptin, ng/ml | -0.4101 | <0.0001 | -0.3221 | <0.0001 | -0.3711 | <0.0001 |

Metabolic risk factors defined by NCEP-ATP III criteria
Values given are $p < 0.01^{**}$ (significant).
r, Correlation coefficient; NCEP, National Cholesterol Education Program; ATP III, Adult Treatment Panel III; WC, waist circumference; WHR, waist to hip ratio; BMI, body mass index; SBP, systolic blood pressure; DBP, diastolic blood pressure; TC, total cholesterol; TG, triglyceride; HDL-C, high density lipoprotein-cholesterol; LDL-C, low density lipoprotein-cholesterol; FPG, fasting plasma glucose; FPI, fasting plasma insulin; HOMA-IR, homeostasis model assessment-insulin resistance

**Figure 2.**
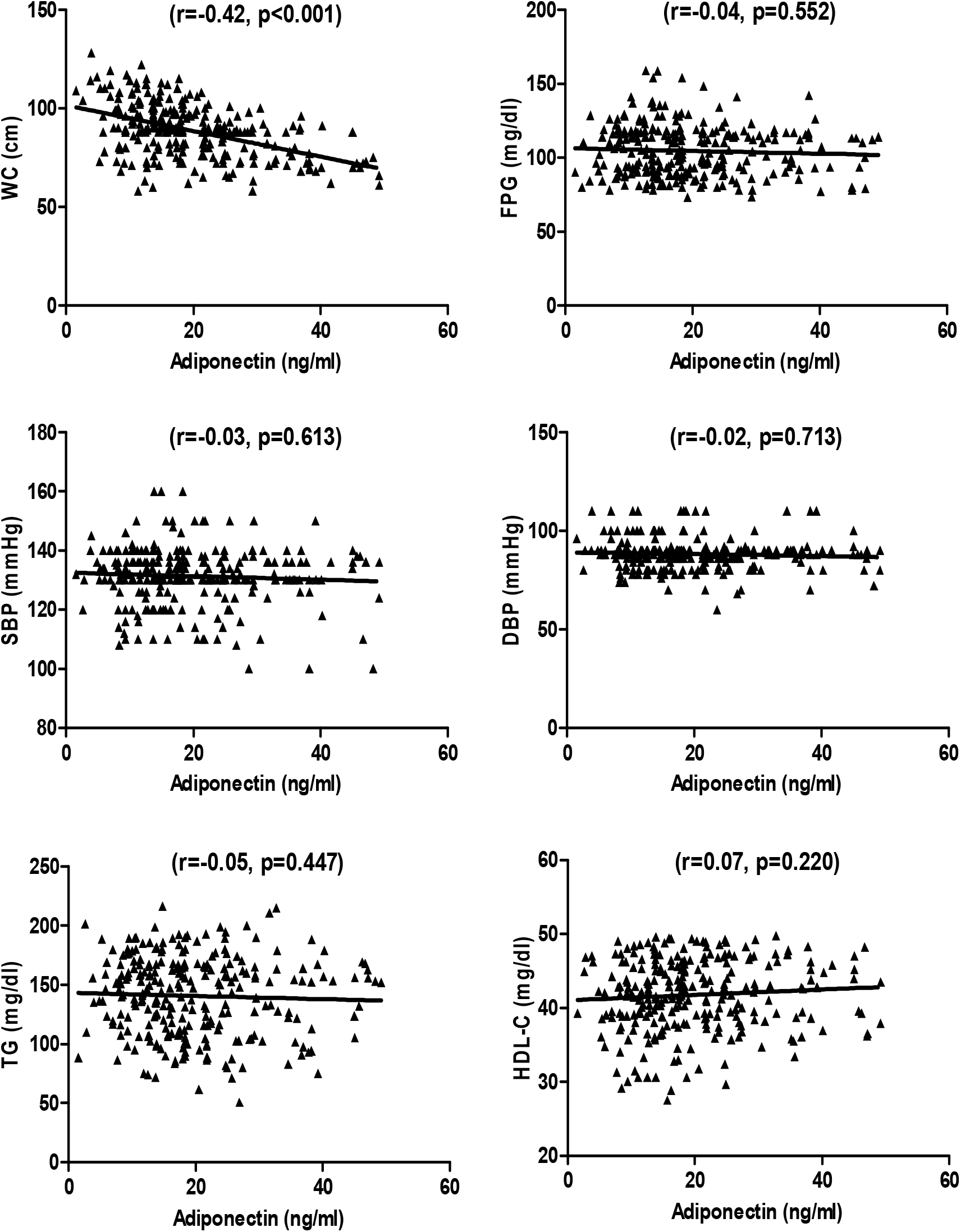
Pearson correlation of serum adiponectin (ng/ml) level with metabolic risk factors in MetS women of North India (n=269). MetS, metabolic syndrome; WC, waist circumference; FPG, fasting plasma glucose; SBP, systolic blood pressure; DBP, diastolic blood pressure; TG, triglyceride; HDL-C, high density lipoprotein-cholesterol

**Figure 3.**
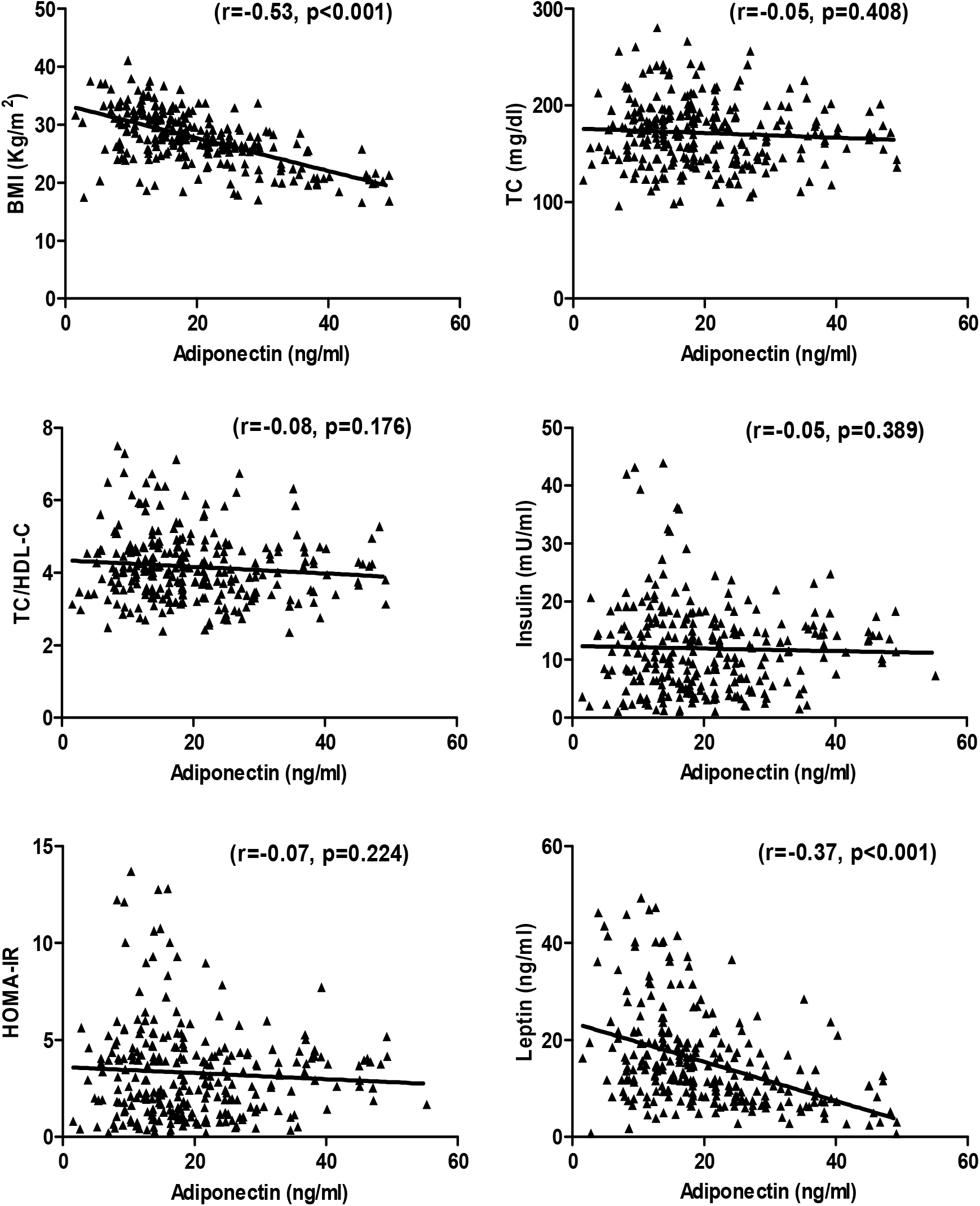
Pearson correlation (r) of serum adiponectin level (ng/ml) with leptin level and other risk factors in MetS women (n=269). MetS, metabolic syndrome; BMI, body mass index; TC, total cholesterol; HDL-C, high density lipoprotein-cholesterol; HOMA-IR, homeostasis model assessment-insulin resistance

## Discussion

The association between adiponectin gene variations and components of MetS, obesity, and HOMA-IR has been extensively studied across various ethnic populations **(Stumvoll M et al., 2002; Yang WS et al., 2006; Mosad AS et al., 2023)**. However, in South India, research on adiponectin and leptin levels is limited. To date, no such studies have specifically investigated the association between adiponectin polymorphisms at loci +45 T/G and +276 G/T with adiponectin and leptin levels, as well as insulin resistance (IR), in adult women with MetS.

In our study, we found that the adiponectin SNPs at +45T/G and +276G/T are significantly associated with higher HOMA-IR and lower adiponectin levels in women with MetS. These particular SNPs were selected based on previous literature and their higher allele frequency in SNP databases. The study also revealed significant differences in serum lipid profiles, including TC, TG, HDL-C, plasma glucose, insulin, HOMA-IR, and serum levels of adiponectin and leptin between women with MetS and those without. Moreover, consistent with previous studies, our findings showed that lower serum adiponectin levels are associated with lower HDL-C or dyslipidemia, obesity, high blood pressure and metabolic syndrome **(Matsubara M et al., 2002; Baratta R et al., 2004; Ryan AS et al., 2003; Kazumi T et al., 2002; Mohan V et al., 2005; Kaur H et al., 2018)**. Collectively, these human studies underscore the importance of adiponectin as a key biomarker for MetS.

A study conducted on white and Pima Indian populations by **Weyer et al. (2001)** found that lower adiponectin levels are more strongly related to the degree of HOMA-IR and hyperinsulinemia than to overall adiposity or glucose intolerance. Evidence suggests that adiponectin plays a significant role in contributing to HOMA-IR and MetS. Due to its insulin-sensitizing properties, adiponectin may influence glucose metabolism by stimulating pancreatic insulin secretion in vivo **(Okamoto M et al., 2008)**. These findings indicate that clinically significant hypo-adiponectinemia is a key feature of MetS, potentially contributing to the increased risk of HOMA-IR. Further studies, particularly in men, are needed to validate these associations.

Adiponectin is often referred to as the “fat-burning molecule” because it helps redirect fatty acids to muscle tissue for oxidation. Like leptin, adiponectin appears to prevent fat deposition in insulin-sensitive tissues by increasing fat oxidation, activating insulin signaling, and upregulating molecules involved in fatty acid transport, oxidation, and energy dissipation, thereby enhancing insulin sensitivity **(Ceddia RB et al., 2005)**. Leptin, a circulating hormone, is responsible for regulating body weight and energy homeostasis. **Fernandez-Real et al. (1997)** found that serum leptin levels are higher in obese individuals compared to those with normal body weight. Similarly, our study revealed that serum leptin levels are higher in women with MetS compared to those wMetS. It is well established that leptin levels are several times higher in women **(Hickey MS et al., 1996)**, suggesting a potentially greater effect on the sympathetic nervous system (SNS) than in men **(Flanagan DE et al., 2007)**. In addition to its role in adiposity, leptin also influences insulin sensitivity and insulin secretion, exerting direct effects on several metabolic actions of insulin and stimulating protein synthesis in insulin-sensitive target cells.

In a genome-wide scan aimed at identifying genetic loci associated with susceptibility to MetS traits, *Kissebah et al*. **(2000)** pinpointed a “hot” region on chromosome 3q27 that overlaps with the adiponectin gene’s location on the human genome. Our findings revealed that the presence of the mutant TG+GG genotype at the +T45G site and the mutant GT+TT genotype at the +G276T site of the adiponectin gene increased the likelihood of having high-risk genotypes by 1.55-fold and 1.40-fold, respectively. Similarly, the mutant G allele at +T45G and the mutant T allele at +G276T were associated with a 1.49-fold and 1.36-fold increased risk, respectively, when comparing MetS and wMetS women. Interestingly, a significant difference in the frequency of the TG+GG genotypes was observed at +45T/G, while the GT+TT genotypes at +276 G/T did not show a significant difference between MetS and wMetS women. These findings suggest that the rate of disease progression varies among individuals, likely due to differences in their genetic susceptibility to MetS.

In our study, we found that polymorphisms at positions +T45G and +G276T of the adiponectin gene may serve as genetic risk factors influencing circulating adiponectin levels in women with MetS. These adiponectin polymorphisms likely contribute to variations in serum adiponectin levels. Our findings are consistent with previous studies that demonstrated a significant association between the +45 T/G and +276 G/T polymorphisms and obesity, insulin sensitivity, and adiponectin levels **(Stumvoll M et al., 2002; Menzaghi C et al., 2002; Fumeron F et al., 2004; Pollin T et al., 2005)** in women with MetS and wMetS. These studies suggest that both SNPs are linked to obesity, HOMA-IR, and serum adiponectin levels. However, the data are not entirely consistent, as some studies have failed to confirm these associations **(Filippi E et al., 2004; Vozarova de Court B et al., 2005)**. *De Luis et al*. **(2016)** also suggested that the +276 variant of adiponectin is more strongly associated with MetS, HOMA-IR, and adiponectin levels.

Our findings also reveal significant associations between anthropometric variables (WC, WHR, BMI, blood pressure), lipid profile (TC, TG, TC/HDL ratio), and biochemical parameters such as fasting insulin concentration and serum leptin levels with the mutant genotypes of the +45 T/G and +276 G/T adiponectin polymorphisms in women with MetS. Notably, no other studies have examined the relationship between these polymorphisms and serum leptin levels in North Indian adult women with MetS. Our study supports the hypothesis that these adiponectin SNPs (+45 T/G and +276 G/T) are directly associated with lower adiponectin levels, higher leptin levels, elevated HOMA-IR, and other metabolic risk factors in women with MetS, particularly within the North Indian population. Consistent with previous research **(Engeli S et al., 2003; Hulthe J et al., 2003**), our findings indicate that circulating adiponectin levels are positively correlated with HDL-C and insulin sensitivity **(Pellme F et al., 2003)** while being negatively correlated with serum leptin levels, HOMA-IR, WHR, adiposity, and MetS **(Hivert MF et al., 2008; Weyer C et al., 2001; Arita Y et al., 1999; Hung J et al., 2008)**.

To the best of our knowledge, this study presents strong evidence of a link between adiponectin gene variants and HOMA-IR and leptin levels in adult women with MetS from North India. These results suggest that these adiponectin variants significantly influence adiponectin and leptin levels, insulin sensitivity, and related risk factors in MetS women of North Indian descent. The limitations of this study include a relatively small sample size and exclusion of male participants of North India, which may affect the generalizability of the findings to other populations or ethnic groups.

In conclusion, our findings suggest that lower adiponectin levels may serve as an independent risk factor and potential biomarker for MetS. Future research could reveal that reduced adiponectin levels specifically signal certain MetS risk factors rather than the entire spectrum currently used for diagnosis. Furthermore, our study indicates that adiponectin polymorphisms at +45T/G and +276G/T might play a protective role in the development of MetS, providing important insights into the genetic underpinnings of the syndrome. However, the influence of other genetic and environmental factors should not be disregarded.

## Data Availability

All data produced in the present study are available upon reasonable request to the authors

## Acknowledgements

We gratefully acknowledge the financial support from the Indian Council of Medical Research, New Delhi (Grant No. 3/1/2/2/06-RHN).

## References

Gupta A, Gupta V. Metabolic syndrome: what are the risks for humans? Biosci Trends. 2010 Oct;4(5):204–12.

Lorenzo C, Serrano-Rios M, Martinez-Larrad MT, Williams K, Gomez-Gerique Stern MP, Haffner SM. Central adiposity determines prevalence differences of the metabolic syndrome. Obesity Res. 2003; 11(12): 1480–487.

Nguyen TMD. Adiponectin: Role in Physiology and Pathophysiology. Int J Prev Med. 2020 Sep 3;11:136. doi: 10.4103/ijpvm.IJPVM_193_20.

Bjørbaek C, Kahn BB. Leptin signaling in the central nervous system and the periphery. Recent Prog Horm Res. 2004;59:305–31. doi: 10.1210/rp.59.1.305.

Mohammadzadeh G, Ghaffari MA, Heibar H, Bazyar M. Association of two Common Single Nucleotide Polymorphisms (+45T/G and +276G/T) of ADIPOQ Gene with Coronary Artery Disease in Type 2 Diabetic Patients. Iran Biomed J. 2016 Jul;20(3):152–60. doi: 10.7508/ibj.2016.03.004. Epub 2016 Jan 19.

Ukkola O, Ravussin E, Jacobson P, Sjöström L, Bouchard C. Mutations in the adiponectin gene in lean and obese subjects from the Swedish obese subjects cohort. Metabolism. 2003 Jul;52(7):881–4. doi: 10.1016/s0026-0495(03)00074-x.

Palit SP, Patel R, Jadeja SD, Rathwa N, Mahajan A, Ramachandran AV, Dhar MK, Sharma S, Begum R. A genetic analysis identifies a haplotype at adiponectin locus: Association with obesity and type 2 diabetes. Sci Rep. 2020 Feb 19;10(1):2904. doi: 10.1038/s41598-020-59845-z. Erratum in: Sci Rep. 2020 Apr 27;10(1):7017. doi: 10.1038/s41598-020-62762-w.

Jang Y, Chae JS, Koh SJ, Hyun YJ, Kim JY, Jeong YJ, Park S, Ahn CM, Lee JH. The influence of the adiponectin gene on adiponectin concentrations and parameters of metabolic syndrome in non-diabetic Korean women. Clin Chim Acta. 2008 May;391(1-2):85-90. doi: 10.1016/j.cca.2008.02.011. Epub 2008 Feb 16.

Expert Panel on Detection, Evaluation, and Treatment of High Blood Cholesterol in Adults. Executive Summary of The Third Report of The National Cholesterol Education Program (NCEP)

Expert Panel on Detection, Evaluation, And Treatment of High Blood Cholesterol In Adults (Adult Treatment Panel III). JAMA. 2001 May 16;285(19):2486–97. doi: 10.1001/jama.285.19.2486.

Matthews DR, Hosker JP, Rudenski AS, Naylor BA, Treacher DF, Turner RC. Homeostasis model assessment: insulin resistance and beta-cell function from fasting plasma glucose and insulin concentrations in man. Diabetologia. 1985 Jul;28(7):412–9. doi: 10.1007/BF00280883.

Stumvoll M, Tschritter O, Fritsche A, Staiger H, Renn W, Weisser M, Machicao F, Häring H. Association of the T-G polymorphism in adiponectin (exon 2) with obesity and insulin sensitivity: interaction with family history of type 2 diabetes. Diabetes. 2002 Jan;51(1):37–41. doi: 10.2337/diabetes.51.1.37.

Yang WS, Chuang LM. Human genetics of adiponectin in the metabolic syndrome. J Mol Med (Berl). 2006 Feb;84(2):112–21. doi: 10.1007/s00109-005-0011-7. Epub 2005 Dec 31.

Mosad AS, Elfadil GA, Gassoum A, Elamin KM, Husain NEOSA. Adiponectin Gene Polymorphisms and Possible Susceptibility to Metabolic Syndrome among the Sudanese Population: A Case-Control Study. Int J Endocrinol. 2023 Apr 27;2023:5527963. doi: 10.1155/2023/5527963.

Matsubara M, Maruoka S, Katayose S. Decreased plasma adiponectin concentrations in women with dyslipidemia. J Clin Endocrinol Metab. 2002 Jun;87(6):2764–9. doi: 10.1210/jcem.87.6.8550.

Baratta R, Amato S, Degano C, Farina MG, Patanè G, Vigneri R, Frittitta L. Adiponectin relationship with lipid metabolism is independent of body fat mass: evidence from both cross-sectional and intervention studies. J Clin Endocrinol Metab. 2004 Jun;89(6):2665–71. doi: 10.1210/jc.2003-031777.

Ryan AS, Berman DM, Nicklas BJ, Sinha M, Gingerich RL, Meneilly GS, Egan JM, Elahi D. Plasma adiponectin and leptin levels, body composition, and glucose utilization in adult women with wide ranges of age and obesity. Diabetes Care. 2003 Aug;26(8):2383–8. doi: 10.2337/diacare.26.8.2383.

Kazumi T, Kawaguchi A, Sakai K, Hirano T, Yoshino G. Young men with high-normal blood pressure have lower serum adiponectin, smaller LDL size, and higher elevated heart rate than those with optimal blood pressure. Diabetes Care. 2002 Jun;25(6):971–6. doi: 10.2337/diacare.25.6.971.

Mohan V, Deepa R, Pradeepa R, Vimaleswaran KS, Mohan A, Velmurugan K, Radha V. Association of low adiponectin levels with the metabolic syndrome--the Chennai Urban Rural Epidemiology Study (CURES-4). Metabolism. 2005 Apr;54(4):476–81. doi: 10.1016/j.metabol.2004.10.016.

Kaur H., Badaruddoza B., Bains V., Kaur A. Genetic association of ADIPOQ gene variants (−3971A>G and +276G>T) with obesity and metabolic syndrome in North Indian Punjabi population. PLoS One. 2018;13(9) doi: 10.1371/journal.pone.0204502.

Weyer C, Funahashi T, Tanaka S, Hotta K, Matsuzawa Y, Pratley RE, Tataranni PA. Hypoadiponectinemia in obesity and type 2 diabetes: close association with insulin resistance and hyperinsulinemia. J Clin Endocrinol Metab. 2001 May;86(5):1930–5. doi: 10.1210/jcem.86.5.7463.

Okamoto M, Ohara-Imaizumi M, Kubota N, Hashimoto S, Eto K, Kanno T, Kubota T, Wakui M, Nagai R, Noda M, Nagamatsu S, Kadowaki T. Adiponectin induces insulin secretion in vitro and in vivo at a low glucose concentration. Diabetologia. 2008 May;51(5):827–35. doi: 10.1007/s00125-008-0944-9. Epub 2008 Mar 28.

Ceddia RB, Somwar R, Maida A, Fang X, Bikopoulos G, Sweeney G. Globular adiponectin increases GLUT4 translocation and glucose uptake but reduces glycogen synthesis in rat skeletal muscle cells. Diabetologia. 2005 Jan;48(1):132–9. doi: 10.1007/s00125-004-1609-y. Epub 2004 Dec 24.

Fernández-Real JM, Gutierrez C, Ricart W, Casamitjana R, Fernández-Castañer M, Vendrell J, Richart C, Soler J. The TNF-alpha gene Nco I polymorphism influences the relationship among insulin resistance, percent body fat, and increased serum leptin levels. Diabetes. 1997 Sep;46(9):1468–72. doi: 10.2337/diab.46.9.1468.

Hickey MS, Israel RG, Gardiner SN, Considine RV, McCammon MR, Tyndall GL, Houmard JA, Marks RH, Caro JF. Gender differences in serum leptin levels in humans. Biochem Mol Med. 1996 Oct;59(1):1–6. doi: 10.1006/bmme.1996.0056.

Flanagan DE, Vaile JC, Petley GW, Phillips DI, Godsland IF, Owens P, Moore VM, Cockington RA, Robinson JS. Gender differences in the relationship between leptin, insulin resistance and the autonomic nervous system. Regul Pept. 2007 Apr 5;140(1-2):37-42. doi: 10.1016/j.regpep.2006.11.009. Epub 2006 Dec 21.

Kissebah AH, Sonnenberg GE, Myklebust J, Goldstein M, Broman K, James RG, Marks JA, Krakower GR, Jacob HJ, Weber J, Martin L, Blangero J, Comuzzie AG. Quantitative trait loci on chromosomes 3 and 17 influence phenotypes of the metabolic syndrome. Proc Natl Acad Sci U S A. 2000 Dec 19;97(26):14478–83. doi: 10.1073/pnas.97.26.14478.

Menzaghi C, Ercolino T, Di Paola R, Berg AH, Warram JH, Scherer PE, Trischitta V, Doria A. A haplotype at the adiponectin locus is associated with obesity and other features of the insulin resistance syndrome. Diabetes. 2002 Jul;51(7):2306–12. doi: 10.2337/diabetes.51.7.2306.

Fumeron F, Aubert R, Siddiq A, Betoulle D, Péan F, Hadjadj S, Tichet J, Wilpart E, Chesnier MC, Balkau B, Froguel P, Marre M; Epidemiologic Data on the Insulin Resistance Syndrome (DESIR) Study Group. Adiponectin gene polymorphisms and adiponectin levels are independently associated with the development of hyperglycemia during a 3-year period: the epidemiologic data on the insulin resistance syndrome prospective study. Diabetes. 2004 Apr;53(4):1150–7. doi: 10.2337/diabetes.53.4.1150.

Pollin TI, Tanner K, O’connell JR, Ott SH, Damcott CM, Shuldiner AR, McLenithan JC, Mitchell BD. Linkage of plasma adiponectin levels to 3q27 explained by association with variation in the APM1 gene. Diabetes. 2005 Jan;54(1):268–74. doi: 10.2337/diabetes.54.1.268.

Filippi E, Sentinelli F, Trischitta V, Romeo S, Arca M, Leonetti F, Di Mario U, Baroni MG. Association of the human adiponectin gene and insulin resistance. Eur J Hum Genet. 2004 Mar;12(3):199–205. doi: 10.1038/sj.ejhg.5201120.

Vozarova de Courten B, Hanson RL, Funahashi T, Lindsay RS, Matsuzawa Y, Tanaka S, Thameem F, Gruber JD, Froguel P, Wolford JK. Common Polymorphisms in the Adiponectin Gene ACDC Are Not Associated With Diabetes in Pima Indians. Diabetes. 2005 Jan;54(1):284–9. doi: 10.2337/diabetes.54.1.284.

de Luis DA, Izaola O, de la Fuente B, Primo D, Fernandez Ovalle H, Romero E. rs1501299 Polymorphism in the Adiponectin Gene and Their Association with Total Adiponectin Levels, Insulin Resistance and Metabolic Syndrome in Obese Subjects. Ann Nutr Metab. 2016; 69(3-4): 226–231. doi: 10.1159/000453401. Epub 2016 Dec 3.

Engeli S, Feldpausch M, Gorzelniak K, Hartwig F, Heintze U, Janke J, Möhlig M, Pfeiffer AF, Luft FC, Sharma AM. Association between adiponectin and mediators of inflammation in obese women. Diabetes. 2003 Apr;52(4):942–7. doi: 10.2337/diabetes.52.4.942.

Hulthe J, Hultén LM, Fagerberg B. Low adipocyte-derived plasma protein adiponectin concentrations are associated with the metabolic syndrome and small dense low-density lipoprotein particles: atherosclerosis and insulin resistance study. Metabolism. 2003 Dec;52(12):1612–4. doi: 10.1016/s0026-0495(03)00313-5.

Pellmé F, Smith U, Funahashi T, Matsuzawa Y, Brekke H, Wiklund O, Taskinen MR, Jansson PA. Circulating adiponectin levels are reduced in nonobese but insulin-resistant first-degree relatives of type 2 diabetic patients. Diabetes. 2003 May;52(5):1182–6. doi: 10.2337/diabetes.52.5.1182.

Hivert MF, Sullivan LM, Fox CS, Nathan DM, D’Agostino RB Sr, Wilson PW, Meigs JB. Associations of adiponectin, resistin, and tumor necrosis factor-alpha with insulin resistance. J Clin Endocrinol Metab. 2008 Aug;93(8):3165–72. doi: 10.1210/jc.2008-0425. Epub 2008 May 20.

Arita Y, Kihara S, Ouchi N, Takahashi M, Maeda K, Miyagawa J, Hotta K, Shimomura I, Nakamura T, Miyaoka K, Kuriyama H, Nishida M, Yamashita S, Okubo K, Matsubara K, Muraguchi M, Ohmoto Y, Funahashi T, Matsuzawa Y. Paradoxical decrease of an adipose-specific protein, adiponectin, in obesity. Biochem Biophys Res Commun. 1999 Apr 2;257(1):79–83. doi: 10.1006/bbrc.1999.0255.

Hung J, McQuillan BM, Thompson PL, Beilby JP. Circulating adiponectin levels associate with inflammatory markers, insulin resistance and metabolic syndrome independent of obesity. Int J Obes (Lond). 2008;32(5):772–9.

